# Nowcasting the COVID–19 pandemic in Bavaria

**DOI:** 10.1101/2020.06.26.20140210

**Authors:** Felix Günther, Andreas Bender, Katharina Katz, Helmut Küchenhoff, Michael Höhle

**Affiliations:** Statistical Consulting Unit StaBLab, Department of Statistics, LMU Munich, Munich, Germany; Department of Genetic Epidemiology, University of Regensburg, Regensburg, Germany; Bavarian Health and Food Safety Authority, Oberschleiβheim, Germany; Department of Mathematics, Stockholm University, Stockholm, Sweden

## Abstract

To assess the current dynamic of an epidemic it is central to collect information on the daily number of newly diseased cases. This is especially important in real-time surveillance, where the aim is to gain situational awareness, e.g., if cases are currently increasing or decreasing. Reporting delays between disease onset and case reporting hamper our ability to understand the dynamic of an epidemic close to now when looking at the number of daily reported cases only. Nowcasting can be used to adjust daily case counts for occurred-but-not-yet-reported events. Here, we present a novel application of nowcasting to data on the current COVID–19 pandemic in Bavaria. It is based on a hierarchical Bayesian model that considers changes in the reporting delay distribution over time and associated with the weekday of reporting. Furthermore, we present a way to estimate the effective time-dependent case reproduction number *R_e_*(*t*) based on predictions of the nowcast. The approaches are based on previously published work, that we considerably extended and adapted to the current task of nowcasting COVID–19 cases. We provide methodological details of the developed approach, illustrate results based on data of the current epidemic, and evaluate the model based on synthetic and retrospective data on COVID–19 in Bavaria. Results of our nowcasting are reported to the Bavarian health authority and published on a webpage on a daily basis (https://corona.stat.uni-muenchen.de/). Code and synthetic data for the analysis is available from https://github.com/FelixGuenther/nc_covid19_bavaria and can be used for adaptions of our approach to different data.

## 1 Introduction

Daily reported case numbers of an infectious disease outbreak do not correspond to the actual number of newly diseased cases on that day. Due to delays from reporting and testing, the number of newly reported cases and the actual number of newly diseased cases can substantially differ. It is the latter, however, that is of central interest when assessing the state and dynamic of an epidemic outbreak. Focusing on the daily number of reported cases hampers our ability to understand current dynamics of the outbreak close to now. This is especially problematic when one wants to gain insight about the current trend or if one want to assess the effects of political and social interventions in real-time. Knowledge of the actual number of new infections per day is highly relevant for the current COVID–19 pandemic, where far-reaching political action was taken in order to contain the epidemic outbreak in 2020.

The problem of occurred-but-not-yet-reported cases during outbreaks is well known from the HIV/AIDS outbreak and different statistical approaches have been proposed to handle delayed reporting. A standard reference is Lawless [1994]. A more flexible Bayesian approach, which is the basis of the model we use here, has been developed by Höhle and an der Heiden [2014]. In the following we will refer to this delay adjustment approach as the *nowcast* and define the reporting delay as the time between *disease onset* and official case reporting by a health authority. Other authors use the term nowcasting for models that focus on adjusting the administrative delay between the first case report to a local health authority and registration (in aggregated data) at higher (e.g., state- and/or federal) authorities [Nicola et al., 2020], or to perform nowcasting of fatal cases between case registration and fatality date [Schneble et al., 2020].

The basic idea of the nowcasting approach proposed here is to estimate the reporting delay between disease onset and reporting date based on observations from the past where both, the disease onset and the reporting date are known. Given the delay distribution and the current number of case reports with reporting dates close to now, we can infer the actual number of new disease onset at current dates. The resulting estimated epidemic curve of newly diseased cases gives a more realistic picture of the current state of the epidemic than looking at daily counts of new case reports. Furthermore, the nowcast can facilitate estimation of the time-dependent, effective reproduction number *R_t_* [Wallinga and Teunis, 2004]. There are other approaches including mathematical infection models (SECIR-model) for the estimation of *R_t_*, see e.g. [Khailaie et al., 2020].

One complication of using nowcasting for COVID–19 reports is that reporting of symptom onset in cases, is not complete: either this information could not be elicited due to difficulties getting in contact with the case or because symptoms had not manifested (yet) at the time of contact with the case. This point was first addressed in Glöckner et al. [2020] and a similar approach based on Lawless [1994] is used by the Robert Koch Institute for analyzing COVID–19 in Germany [an der Heiden and Hamouda, 2020].

Using our approach, we provide nowcast estimates for the COVID–19 pandemic in Bavaria using data from the Bavarian Health and Food Safety Authority (LGL) including the estimation of *R_t_*. The results are updated daily with recent data. In this manuscript, we provide methodological details, show results based on data obtained from the LGL until April, 9th 2020, 10am, and provide results of the evaluation of the proposed nowcasting approach.

## 2 Data

We use daily data on reported COVID–19 cases from Bavaria from the mandatory notification data based on the German Infection Protection Act (IfSG). The data is provided by the Bavarian Health and Food Safety Authority (LGL) on a daily basis and includes a list of all reported cases with the date of reporting to the LGL, the date of reporting to the local health authority (*Gesundheitsamt*), the date of disease onset and the district of residence for the case (*Kreis*). Since we get our data from the LGL, the number of cases reported to the LGL on a specific date is complete and will not change on subsequent days. This consistent data offers a valid base for inferring the epidemic curve and the considered associated quantities.

The date of reporting to the local health authority is closer to disease onset due to a delay between reporting at the local health authority and transmission to the LGL. However, based on the data obtained from the LGL, the aggregated number of cases reported to the local health authorities on a given day may be incomplete because a case reported to the local health authority can be reported to the LGL with a delay of several days and therefore may not be included in the dataset yet. Therefore, we use the date of reporting to the local health authority only for the imputation of missing disease onsets, while the nowcast is based on the date a case was reported to the LGL (cf. Steps 1 and 2 in Section 3.1).

Information on disease onset stems from a retrospective collection of the day of symptom onset. However, the daily COVID–19 surveillance data of Bavaria contain, about 50%-60% cases with missing information on day of symptom onset in the weeks close to now. For a specific week, this fraction becomes lower over time since more information on the cases is collected. The missing onset information exists partly due to the heavy workload imposed on health authorities during the pandemic, but also because a certain proportion of cases have no or only very mild symptoms. However, we expect the later explanation to be less prominent that the former.

Note also that the date of symptom onset does not correspond to the infection date due to a preceding incubation time.

## 3 Methods

In the following sections, we provide methodological details regarding the proposed nowcasting (cf. Section 3.1 as well as the estimation of the time-dependent case reproduction number (cf. 3.2). The nowcast itself consists of two steps: Imputation of missing disease onset dates (Step 1) and Bayesian nowcasting based on the imputed data (Step 2).

### 3.1 Nowcasting

Due to the many cases with a missing disease onset date, we decided to proceed with a two-step approach for nowcasting. Firstly, we impute missing data on disease onset and, secondly, perform the nowcast based on the information on reporting date (available for all cases) and the date of disease onset, which is partly available and partly imputed. Imputing missing disease onset information implies that we also consider presymptomatic and asymptomatic COVID–19 cases in our analyses (to the part at which they are observed in the official COVID–19 case counts). The rationale is, that this allows to compare the nowcasting results to the daily updated reported case numbers. Furthermore, focusing on symptomatic cases only is problematic since for cases with missing disease onset it is not completely clear whether they are asymptomatic, just symptomless at the date of reporting (pre-symptomatic), or actually show symptoms but information on symptom onset is missing for other reasons, e.g., not yet collected.

Step 1: Imputation of disease onset In the imputation step, we fit a flexible generalized additive model for location, scale and shape [GAMLSS, Stasinopoulos et al., 2017], assuming a Weibull-distribution for the delay time *t_d_* > 0 between disease onset and reporting date at the local health authority:

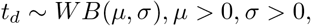

where *μ* and *σ* are the location and scale parameters of the Weibull distribution with density 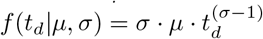. The same, additive predictor (1) was defined for both, *μ* and *σ*

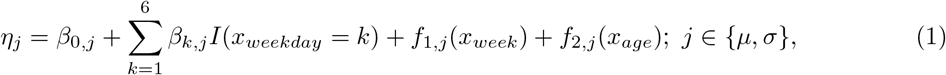

however, the estimated effects could differ for the two distributional parameters. In (1), parameter *β*_0_,*_j_* is the location- or scale-specific global intercept and *β_k_*,*_j_* is the effect of the weekday on which the report arrived at the local health authority. Furthermore, *f*_1_,*_j_* and *f*_2,_*_j_* are smooth effects of the calendar week (of report arrival) and age of case, respectively, both parameterised via cubic splines.

To estimate the model we use data of all cases for which the disease onset date and thereby *t_d_* is available. Afterwards, we impute the delay time *t_d_*, if missing, by sampling from the fitted, conditional Weibull distribution and derive the missing symptom/disease onset date. No imputation is performed for observations for which the symptom onset date is reported.

Since this imputation induces, conditional on the predictors of the GAMLSS imputation model, a missing at random assumption with respect to the time between disease onset and case reporting, we perform a sensitivity analysis, where we omit (i) all individuals where the reports say explicitly that they were symptom-free and (ii) all individuals with missing information about symptoms. This allows us to check, whether the dynamics of the daily number of individuals with available symptoms are structurally different compared to all registered cases over time.

Step 2: Bayesian nowcasting For the nowcasting step, we use a Bayesian hierarchical model based on Höhle and an der Heiden [2014] which associated implementation in the R-package surveillance [Salmon et al., 2016]. In the present work, we have extended the approach considerably, adapted it to the context of COVID–19, and provide a novel implementation in rstan [Stan Development Team, 2020].

Let *N_t,d_* = *n_t,d_* be the (observed) number of cases, with disease onset on day t and reported with a delay of d days (case report arrives on day *t* + *d*). On day *T* > *t* (“current” day, i.e. “now”), the information is available on 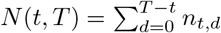 cases that had disease onset on day *t* and are reported until day *T*. The aim of nowcasting is to predict the unobserved total number of newly diseased cases on day *t*, *N*(*t*, ∞) 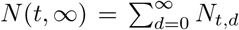, based on information available up until the current day *T*. For identifiability reasons, one defines a maximum relevant delay time of *d* = *D* and considers each observation with an observed delay > *D* as having a delay of *D*. As described in Höhle and an der Heiden [2014], the hierarchical Bayesian model for nowcasting consists essentially of two parts: a model for the expected number of newly diseased cases on day *t*, *E*(*N*(*t*, ∞)) = *λ_t_*, and a model for the delay distribution at day t, specifying the probability of a reporting delay of d days for a case with disease onset at day *t*, *P*(delay = *d*|onset = *t*) = *p_t,d_*. Both parts of the model can in general be flexibly specified. We set the maximum delay to *D* = 21 and utilize the following hierarchical model for nowcasting:

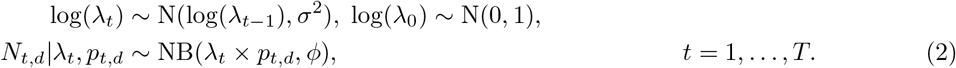

The number of cases with disease onset at day *t* and reporting delay *d* days, *N_t,d_*, is assumed to follow a Negative Binomial distribution with expectation *λ_t_* × *p_t,d_*, and overdispersion parameter *ϕ*. For the delay distribution we utilize a discrete time hazard model *h_t,d_* = *P*(delay = *d*|delay ≥ *d*, *W_t,d_*) as

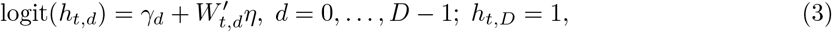

where *W_t,d_* is a vector of time- and delay-specific covariates and *η* the corresponding regression coefficients. In our main model, we use linear effects of time with breakpoints every two weeks before the current day (corresponding to a first order spline), and a categorical weekday effect of the reporting day with a common effect for holidays and Sunday, since there are substantial differences in the reported case numbers over the week. From model (3), we can derive the probabilities of interest in (2), *p_t_*_,0_ = *h_t_*_,0_ and 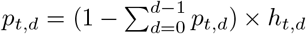. The main goal of of nowcasting is to obtain inference about 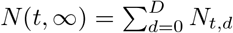. Based on the described Bayesian hierarchical model, this corresponds to a sum of Negative Binomial distributed counts and we can obtain such inference by summing up the MCMC samples of *N_t,d_* at each timepoint *t*. In an alternative specification of the model during evaluation (see below) we assume that *N_t,d_*|*λ_t_*,*p_t,d_* — Po(*λ_t_* × *p_t,d_*). In this case *N*(*t*, *∞*) is Poisson distributed as well and it is directly possible to sample from Po(*λ_t_*) to perform inference about *N*(*t*, *∞*).

The utilization of the first-order random walk for modeling *λ_t_* in (2) was motivated by results of McGough et al. [2020]. For the modeling of the delay distribution we utilized several different approaches and covariates and evaluated them on synthetic data and retrospectively on the Bavarian COVID–19 data (see below for a description of the approaches).

### 3.2 Estimation of the time-dependent case reproduction number *R_e_*(*t*)

Once a depletion of susceptibles occurs during an outbreak of a person-to-person transmitted disease or specific interventions are made, a key parameter to track is the so called effective reproduction number (also refered to as case reproduction number). This time-varying quantity is defined as follows: Consider a case with disease onset on day *t* - the expected number of secondary cases one such primary case generates will be denoted by *R_e_*(*t*). The time until these secondary cases will show symptoms is governed by the serial-interval distribution, which is defined as the time period between manifestation of symptoms in the primary case to time of symptom manifestation in the secondary case [Svensson, Å., 2007].

We estimate the time-dependent case reproduction number by the procedure of Wallinga and Teunis [2004]: Consider a case *j* showing symptoms for the first time on day *t_j_*. The relative likelihood that a case *i* (with symptom onset on day *t_i_*) was infected by *j* is given by

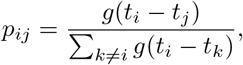

where *g* is the PMF of the serial-interval distribution. For the serial interval distribution we use a discretized version of the results from Nishiura et al. [2020], which find a log-normal distribution with mean 4.7 days and standard deviation 2.9 as the most suitable fit to data from 28 infector-infectee pairs. An estimate of the effective reproduction number at time *t* is now given as the average reproduction number of each case *j* showing first symptoms of the illness on day *t*:

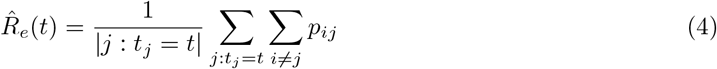

We prefer this *R_e_*(*t*) estimation over the method used in an der Heiden and Hamouda [2020], because it is unbiased for our generation time distribution (see the discussion in Höhle [2020]). For each imputed dataset, we extract *K* = 500 time series of case counts from the posterior distribution of the nowcast and then estimate *R_e_*(*t*) as defined in (4) for each time series using the R-package R0 [Obadia et al., 2012]. Furthermore, each *R_e_*(*t*) estimation generates *M* = 100 samples from the corresponding sampling distribution of *R_e_*(*t*). Altogether, we report 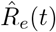 as mean of these *K* × *M* samples together with the 2.5% and 97.5% quantiles to form a 95% credibility interval for *R_e_*(*t*). We estimate *R_e_*(*t*) for all t so that *t* + *q_g_*(0.95) ≤ *T*, where *q_g_*(0.95) is the 95% quantile of the serial interval distribution. This avoids a downward bias in the *R_e_*(*t*) estimation near “now”. Alternatively, one could employ correction methods near *T* [Cauchemez et al., 2006].

### 3.3 Evaluation of the methods

We perform an evaluation of the hierarchical nowcasting based on synthetic data mimicking the reported Bavarian COVID–19 data and retrospectively on the official data from the LGL that was reported until July, 31. For creation of the synthetic data, we utilized a smoothed version of the observed number of reported disease onsets per day and specified a reporting delay model similar to the model described in (3) with five changepoints in the linear time effect on the hazard. This leads firstly to a slight increase, followed by a decrease and stabilization, and a final slight increase of the (average) reporting delay over time (see **Supplemental Note** for a detailed description and visualisation of the data generating process). The aggregated daily numbers of disease onsets and daily numbers of reported cases are similar in structure to the officially reported data. For faster computation during the evaluation, we divided the daily cases by two. For the retrospective evaluation on the official COVID–19 data we focus on all reported cases with available disease onset and on the time period between March, 17 and June, 30, assuming that all cases that will be reported with disease onset until June, 30 are reported on July, 31.

For the evaluation of the nowcasting we estimate several different models (Table 1). We vary the distributional assumptions of *N_t,d_* between Poisson and Negative Binomial (cf. Section 3.1, Step 2). Furthermore, we vary the specification of the model for the reporting delay distribution: firstly, we assume a reporting delay distribution without changes over time, secondly, we estimate linear effects of time on the delay distribution with changepoints every two weeks, and thirdly use a different specification of the discrete time hazard model, where we model

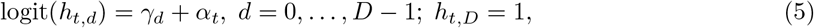

with a prior on 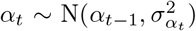 and *α*_0_ = 0. With this model, we aim to estimate smooth daily changes in the delay distibution over time similar to the first-order random walk in modeling of *λ_t_*. In case of the synthetic data, we additionally estimate the nowcasting with the known true changepoints in the delay distribution (that are unknown in real world applications) and in case of the retrospective evaluation on Bavarian data, we additionally include in some scenarios dummy effects of the weekday of the reporting date.

**Table 1:** Estimated hierarchical nowcast models in the evaluation on synthetic and actual Bavarian COVID–19 data.

|  |  |
| --- | --- |
| Synthetic data |  |
| Distribution $N_{t,d}$ | Delay distribution |
| Poisson | No changes |
| Poisson | Linear time-effect with changepoints every two weeks |
| Poisson | Linear time-effect with true changepoints |
| Negative Binomial | Linear time-effect with changepoints every two weeks |
| Negative Binomial | Linear time-effect with true changepoints |
| Negative Binomial | Daily changes (first order random walk) |
| Retrospective evaluation on Bavarian data |  |
| Distribution $N_{t,d}$ | Delay distribution |
| Poisson | No changes |
| Poisson | Linear time-effect with changepoints every two weeks |
| Negative Binomial | Linear time-effect with changepoints every two weeks |
| Negative Binomial | Daily changes (first order random walk) |
| Negative Binomial | Linear time-effect with changepoints every two weeks + reporting weekday effect |
| Negative Binomial | Daily changes (first order random walk) + reporting weekday effect |

To compare the performance of the different models, we estimate the log scoring rule (logS) and the continuous ranked probability score (CRPS) [Jordan et al., 2019], root mean squared error (RMSE), as well as coverage frequencies of 95-% prediction intervals. For all those criteria, we average over all dates and nowcast predictions 2-6 days before the current date. In addition to the quantitative measures, we visually inspect the performance of the different approaches based on the nowcasting predictions and the estimated delay distribution in comparison to the empirical truth in order to identify potential problems of the models.

We extend the retrospective evaluation of the nowcasting on Bavarian data to the estimation of R_e_(t) and compare the estimated 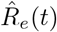 on the most current day max(*t*) s.t. *t* + *q_g_*(0.95) ≤ *T* for all *T* to the retrospective *true R_e_*(*t*) given all available case data until July, 31. This is done based on all evaluated models, and we visually inspect the estimated 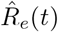 over time and compute coverage frequencies of 95-% credibility intervals.

### 3.4 Implementation

All calculations were done using the statistical programming environment R [R Core Team, 2020]. Nowcasting was performed based on a custom rstan [Stan Development Team, 2020] implementation. Estimation of *R_e_*(*t*) was based on code of the R0 package [Obadia et al., 2012] for each MCMC sample. For computation of the proper scoring rules we used the scoringRules package [Jordan et al., 2019]. Code to reproduce our analysis and for adaption to other application scenarios is available at https://github.com/FelixGuenther/nc_covid19_bavaria. There, we also provide an artificial dataset based on the observed reporting dates of cases but for data protection reasons featuring only artificial information on the age and disease onset dates of the cases.

## 4 Results

### 4.1 Data

We present results based on data obtained from LGL on April 9, 2020, 10am. The data contain information on 29,262 COVID–19 cases which we restrict to 29,246 cases reported after 2020-03-01, as the first 16 COVID–19 cases reported between 2020-01-28 and 2020-02-13 (reported disease onset between 2020-01-23 and 2020-02-03, 3 with missing onset information) concerned a contained outbreak [Böhmer et al., 2020] and no further cases were detected upon 27.02.2020. This outbreak can therefore be assumed to not have contributed to the later disease spread.

Information on disease onset is available for 13,137 cases, but reported disease onset was past the official reporting date for 50 cases and before 2020-01-23 for 16. We set the disease onset date for these cases as missing, yielding 13,071 cases with valid information on disease onset (44.7%). For these, the median delay between disease onset and reporting was 7 days (25%-quantile: 5, 75%-quantile: 11), Table 2 shows observed delay times over the observation period and reveals a considerable increase in the delay distribution over time.

**Table 2:** Week-specific observed number of cases with available information on disease onset. Empirical mean, median and 25%-/75%-quantile of distribution of delay between disease onset and reporting at local health authority. Cases are grouped into weeks based on their reporting date at local health authority. Data from April, 9, 10am.

| Rep. Week | n | Delay available | % avail. | Mean | Median | 25%-Quant. | 75%-Quant. |
| --- | --- | --- | --- | --- | --- | --- | --- |
| 10 | 114 | 77 | 68% | 5.8 | 5 | 4 | 8 |
| 11 | 1074 | 459 | 43% | 5.4 | 5 | 3 | 7 |
| 12 | 4660 | 2100 | 45% | 6.0 | 5 | 4 | 8 |
| 13 | 8858 | 4268 | 48% | 7.5 | 7 | 4 | 10 |
| 14 | 11003 | 4800 | 44% | 8.8 | 8 | 5 | 12 |
| 15 | 3532 | 1335 | 38% | 8.9 | 7 | 4 | 12 |

### 4.2 Imputation of missing disease onset

For imputation of missing disease onset dates we estimate a Weibull GAMLSS with smooth effects of the reporting week, the cases’ age, and a categorical effect of the weekday of report arrival on location and scale. We utilize the reporting date at the local health authority in the imputation model, since it is closer to the actual disease onset than the reporting date at LGL and is available for all cases contained in our data. Thereby, we do not have to deal with the additional reporting delay between the local health authorities and the LGL which might also change over time for the imputation. Figure 1 shows the estimated association of the covariates with the median delay. All covariates turned out to be relevant: we find an increase in expected delay time over the reporting weeks, lower reporting delay for older cases and differences over the course of a week. The estimated GAMLSS model is used to impute the date of disease onset for cases with missing onset information.

**Figure 1:**
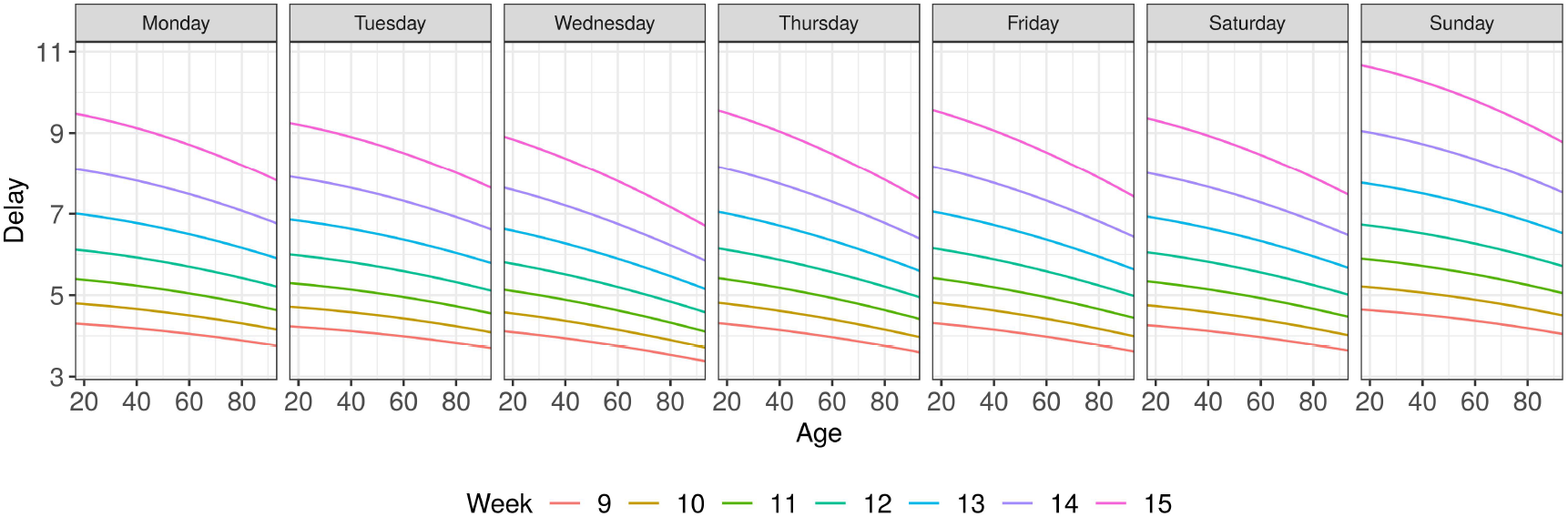
Results of the Weibull GAMLSS imputation model. Shown is the estimated median of the delay time given case-specific covariates (reporting week, weekday of reporting, age).

### 4.3 Nowcasting

Figure 2 shows the number of daily reported cases and the number of cases with reported and imputed disease onset on a certain day over time. Furthermore, we display the estimated new cases from nowcasting based on our main model (2). We observe a clear difference between the estimated new cases from the nowcast and the daily numbers of reported cases. The induced bias due to the reporting delay is obvious: the estimated daily new cases stabilize from around March, 20 on and start to decrease aferwards, while the reported cases still show a rapid increase. The 95%-prediction interval, however, shows substantial uncertainty in estimates, especially for more recent estimates. Note that we set the current day for the nowcasts to April 8th, 2020, since we only consider days with fully available reporting data. Furthermore, we set a reporting lag between the current date and reported nowcast results of two days due to considerable uncertainty in the nowcasts for dates with very few observations with reported or imputed disease onset.

**Figure 2:**
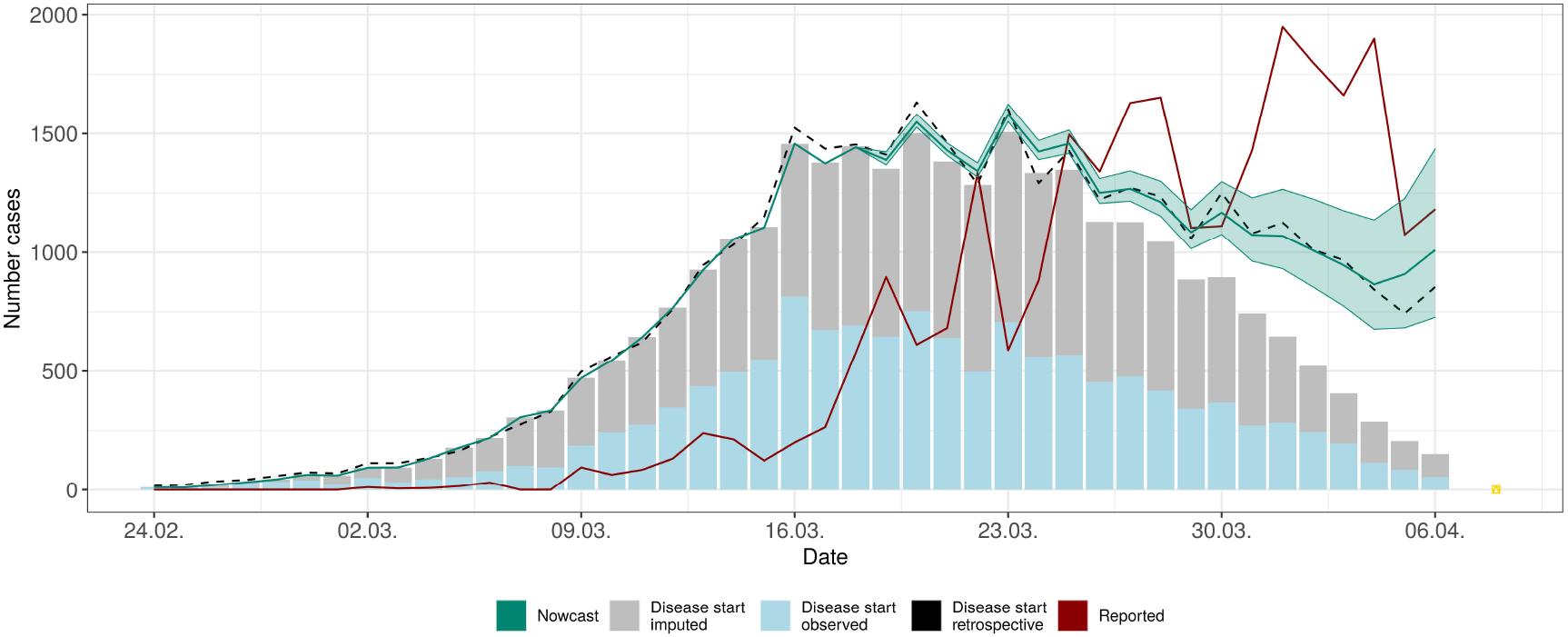
Nowcasting based on Bavarian COVID–19 data until April, 8th, 2020. Shown is the point estimate + 95%-prediction interval of the daily number of newly diseased cases on a given day based on Bayesian hierarchical nowcast. The model considers changes in delay distribution over time based on a linear time effect with two-week changepoints and effects of the weekday of reporting. The expected number of new cases is modeled based on a first order random walk. Additionally, we show the observed number of cases with disease onset (reported and imputed) that are known at April, 8, based on daily bars, the number of newly reported cases per day (red line), and the retrospective *true* number of disease onsets known up until July, 31 (black dotted line). The current day for nowcasting is April, 8, and nowcasts are performed up until April, 6.

The black dotted line in 2 shows the retrospective *true* number of disease onsets (reported and imputed) based on data known at July, 31. We can see that the predicted number of new cases per day and the actually observed number of cases match closely and the prediction intervals contain the actual number of onsets for most days. Note, that the imputation of missing disease onset dates was performed based on the same Weibull GAMLSS but based on different data (all data available at April, 8 and July, 31, respectively), and the number of cases with imputed disease onset on a specific date can therefore vary slightly.

### 4.4 Estimation of the time-dependent case reproduction number

Figure 3 depicts the estimated *R_e_*(*t*) as defined in (4) for the time frame from 24th of February until the 27th of March. This time range is defined by the time of the first secondary case observed in the data and the date of the nowcast minus the number of days it takes for 95% of secondary cases to be observed, which is determined based on 95% quantile of the assumed generation time distribution (10 days). According to the estimate, *R_e_*(*t*) decreased steadily since the beginning of the outbreak and is about *R_e_*(*t*) = 1 since March 20, with *R_e_*(*t*) = 0.81 (CI = (0.75,0.87)) on March 27. However, care is required, if interpreting this result with the timing of interventions, because the *R_e_*(*t*) estimator is defined forward in time and describes the infection process within the following 10 days.

**Figure 3:**
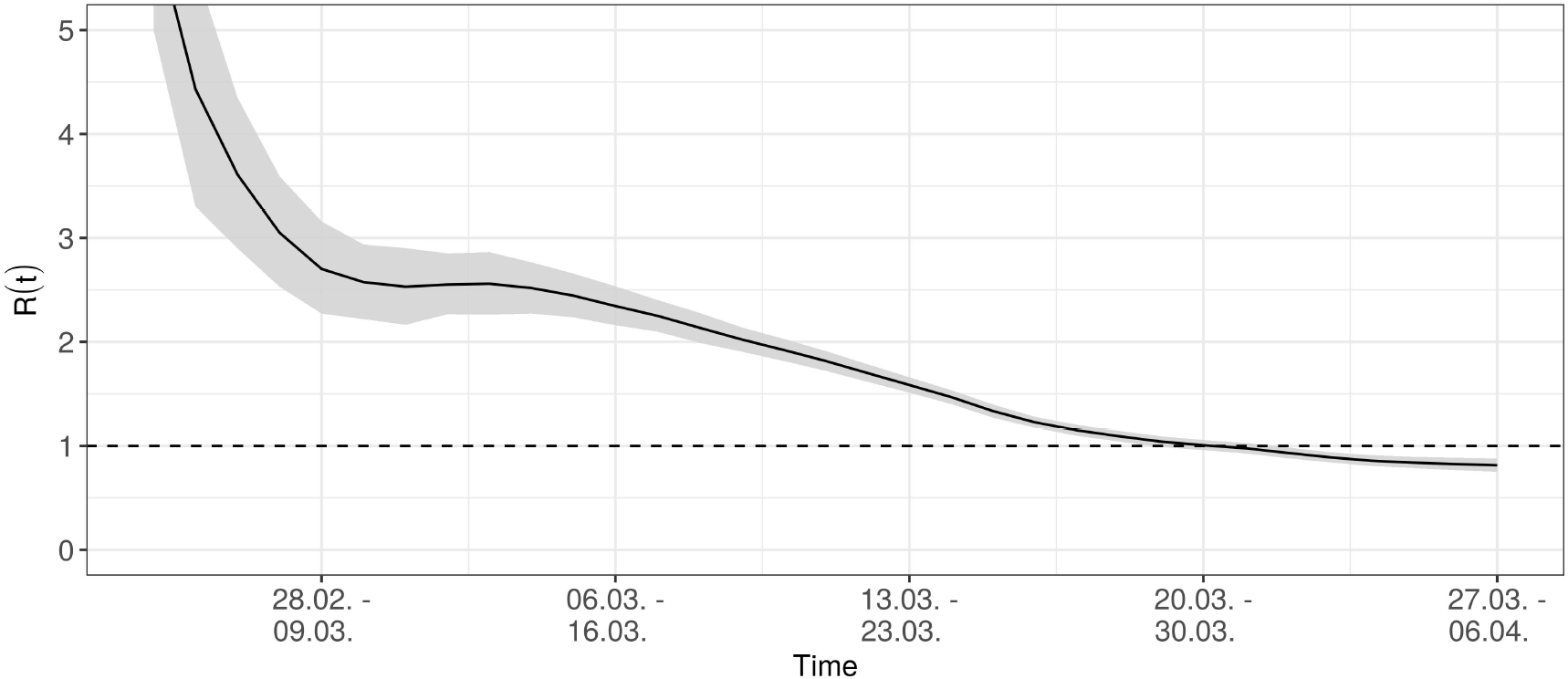
Estimated, time-dependent effective case reproduction number *R_e_*(*t*).

**Figure 4:**
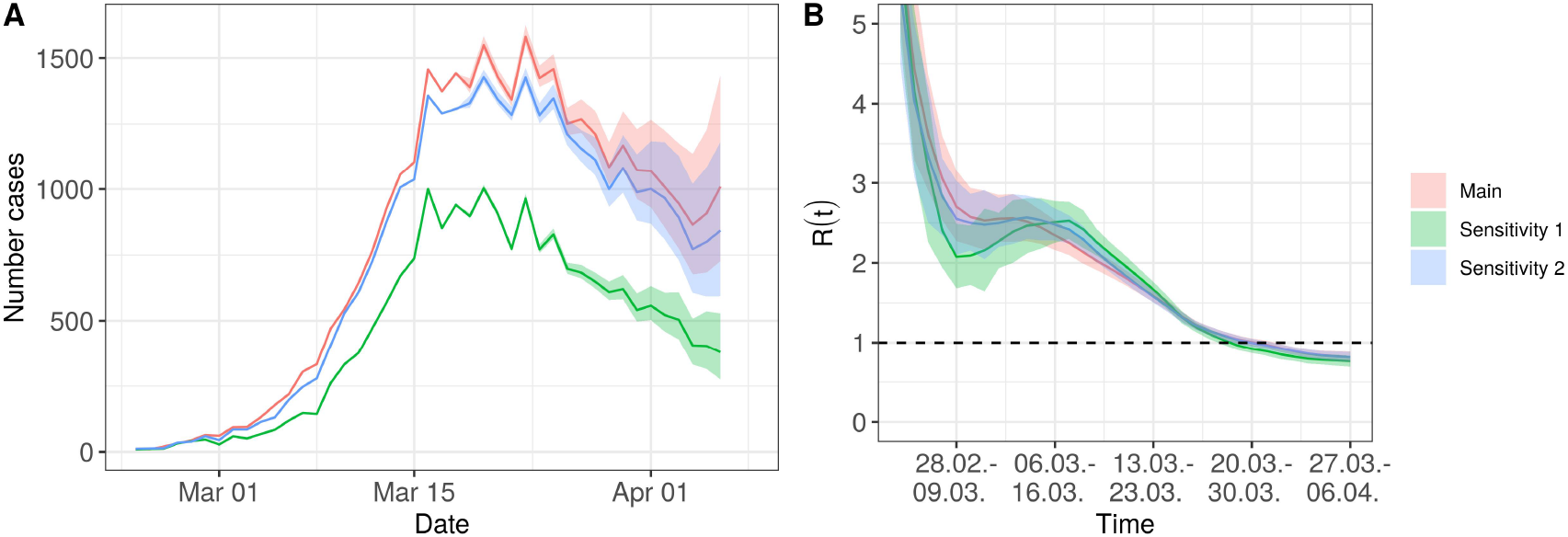
Sensitivity analysis. Estimation of nowcast and *R_e_*(*t*) with associated 95% prediction and credibility intervals based on three different data subsets: original analysis (red), analysis only with cases with reported COVID–19 symptoms (green) and with all cases excluded that are reported as having no symptoms (blue).

### 4.5 Evaluation of nowcast and estimation of *R_e_*(*t*)

We performed an evaluation of the nowcasting approach based on synthetic data and retrospectively on the Bavarian COVID–19 data to investigate the performance of the Bayesian hierarchical model under various model specifications and gain a better understanding of important aspects of modeling. For the synthetic data we found the following (Table 3, more detailed results in **Supplemental Note**): when we supply the true, in reality unknown changepoints of the delay distribution to model fitting the nowcasting approach performs best with respect to our evaluation metrics. Averaged over all days *T*, and for all nowcast days *t* = *T* − 6, … *T* − 2 it shows the lowest log and CRPS score, lowest root mean squared error and shows the desired coverage frequencies for the 95%—prediction intervals. With the models assuming changepoints in the linear time effect on reporting delay every two weeks before *T*, we obtain similar, but slightly worse performance (see **Supplemental Note** for more details). The approach appears to be able to capture moderate changes in the delay distribution successfully. Modeling the changes on a daily basis shows a slightly worse performance with respect to the CRPS score and PI coverage frequencies. Assuming a constant reporting delay distribution over time and ignoring the changes leads to the worst performance with biggest scores and low coverage of the prediction intervals. When specifying an adequate model for the delay distribution, the distributional assumptions regarding *N_t,d_* play a minor role for the evaluation based on synthetic data.

**Table 3:** Results of the evaluation of different nowcasting models on synthetic and actual Bavarian data (retrospectively). logS denotes the log scoring rule, CRPS is the continuous ranked probability score, RMSE denotes the root mean squared error of the posterior median. Additionally we provide coverage frequencies of 95% prediction intervals for the number of newly diseased cases per day and coverage frequencies of 95% credibility intervals in the estimation of *R_e_*(*t*). All scores are averaged over nowcasts for *T* − 6,…, *T* − 2 days, with *T* from March, 17 to June, 30. For R(t), we compute coverage frequencies frequencies for the estimate closest to the current date *T* over all *T*’s.

| Synthetic data |  |  |  |  |  |  |
| --- | --- | --- | --- | --- | --- | --- |
| Model |  | CRPS | logS | RMSE | Cov. 95%-PIs |  |
| Distr. $N_{t,d}$ | Delay model | | | | $N(t, \infty)$ | |
| Poisson | No changes | 46.68 | 13.24 | 89.75 | 0.53 |  |
| Poisson | Lin. effect of time<br>changepoints two weeks | 12.53 | 3.68 | 36.22 | 0.95 |  |
| Neg. Binomial | Lin. effect of time<br>changepoints two weeks | 12.47 | 3.68 | 36.01 | 0.95 |  |
| Neg. Binomial | Daily changes (first order RW) | 28.37 | 3.90 | 92.33 | 0.91 |  |
| Poisson | Lin. effect of time<br>true changepoints | 11.88 | 3.63 | 35.31 | 0.95 |  |
| Neg. Binomial | Lin. effect of time<br>true changepoints | 11.90 | 3.62 | 35.48 | 0.96 |  |
| Retrospective Bavarian COVID-19 data |  |  |  |  |  |  |
| Model |  | CRPS | logS | RMSE | Cov. 95%-PIs/CIs |  |
| Distr. $N_{t,d}$ | Delay model | | | | $N(t, \infty)$ | $R_e(t)$ |
| Poisson | No changes | 193.43 | Inf | 389.56 | 0.19 | 0.56 |
| Poisson | Lin. effect of time<br>changepoints two weeks | 74.32 | Inf | 226.90 | 0.67 | 0.86 |
| Neg. Binomial | Lin. effect of time<br>changepoints 2 weeks | 61.79 | 5.05 | 205.59 | 0.84 | 0.92 |
| Neg. Binomial | Daily changes (first order RW) | 79.21 | 4.83 | 274.70 | 0.86 | 0.95 |
| Neg. Binomial | Lin. effect of time<br>changepoints 2 weeks + weekday effect | 56.63 | 5.22 | 170.59 | 0.82 | 0.92 |
| Neg. Binomial | Daily changes (first order RW)<br>+ weekday effect | 67.32 | 4.99 | 236.05 | 0.90 | 0.94 |

In the retrospective evaluation of the Bavarian data the Poisson model assuming no changes in the reporting delay distribution performs badly as well. This is in line with the apparent changes in the reporting delay between disease onset and reporting at LGL over time (**Supplemental Note**). Comparing the Poisson model with two-week changepoints with a similar model using a Negative Binomial distribution for *N_t,d_* we find the latter to perform better. Adding weekday effects to the delay distribution improves the performance of the models as well. Comparing the Negative Binomial model with daily changes in the delay distribution with the two week changepoint model, we found better coverage frequencies for the former (e.g. 90% vs. 82% when including the weekday effect) but lower CRPS score and RMSE for the latter. Comparing the estimated 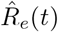 at most current t’s based on the different nowcast models with the retrospective *truth* based on all reported data, we find coverage probabilities of the 95%— credibility intervals bigger than 90% for all Negative Binomial models that consider changes in the delay distribution over time. The estimation of *R_e_*(*t*) is, however, biased when it is based on a biased nowcasting approach, e.g., when changes in the delay distribution are ignored.

## 5 Discussion

Our analyses show that nowcasting is a valuable real-time tool to gain situational awareness in the middle of an outbreak. Based on our evaluation, we found several aspects to be important for the successful application of nowcasts: First and foremost, it is important to account for existing changes in delay between disease onset and case reporting over time. Ignoring such changes can severely bias the predicted number of newly diseased cases. In the Bavarian data, we also found evidence for changes in reporting delay associated with the weekday of reporting which should be accounted for. Secondly, we found an improved performance when modeling the daily counts of newly diseased cases with a specific delay *d*, *N_t,d_*, based on a Negative Binomial distribution with overdispersion. In our data, the case counts show bigger variability then implied by a Poisson distribution. Thirdly, utilizing a first-order random walk for modeling the logarithmic expected daily number of newly diseased cases, *λ_t_*, as proposed by McGough et al. [2020], worked well. We also tried i.i.d. log-Gamma priors and a smooth modeling of the epidemic curve based on truncated power splines as proposed in Höhle and an der Heiden [2014], but found the first-order random walk to perform best. Altogether, we found that a Negative Binomial model with random-walk prior of *λ_t_* and modeling of the delay distribution via an discrete time hazard model with linear time effects and 2 week changepoints works satisfactory. With this model, we are able to account flexibly for changes in reporting delay over time and obtain a satisfactory performance on synthetic data as well as the true retrospective Bavarian COVID–19 data. The alternative smooth modeling of the delay distribution based on daily changes using a first order random walk performed well for many days as well but had convergence issues on some dates and might be overly complex for many scenarios.

However, there are important limitations of any nowcasting estimation: (i) we correct for a bias due to delays between disease onset and case reporting, but provide no correction for possible cases in the population that were not tested. This is a big issue in understanding COVID–19 spread, since there are possibly many undetected cases. Assuming a constant factor of under-reporting we can analyze the dynamics of the outbreak in a more reliable way by our nowcasting method compared to focusing on daily counts of newly reported cases. Furthermore, *R*(*t*) estimates would be invariant to such constant under-reporting. However, if the proportion of undetected cases varies over time, then the dynamics of the epidemic is not described adequately by our approach as well. (ii) We model the temporal variation in the delay distribution in a flexible way. However, short term changes, especially in the time close to the current day can lead to a bias, because it is particularly hard to distinguish between developments in the epidemic curve and changes in the reporting delay with no or very little data. (iii) Our imputation method includes a missing at random assumption, which implies that the time between disease onset and reporting is the same for individuals with and without available symptom onset date. This could be violated due to many asymptomatic and pre-symptomatic among the reported COVID–19 cases. However, the sensitivity analyses in the appendix (5) show that our results are relatively stable to variations of this definition.

Comparing our approach to the one used by the Robert Koch Institute an der Heiden and Hamouda [2020] we use a more detailed delay distribution modeling for the nowcast, e.g., including the day of the week in our model, which turned out to be relevant in our data. Furthermore, we observed and modeled a dependence of the delay time on calendar time as part of the nowcast. This was not originally taken into account by an der Heiden and Hamouda [2020]. When calculating the effective reproduction number *R_e_*(*t*), an der Heiden and Hamouda [2020] used a constant generation time of four days, while our approach includes a more realistic assumption of an individually varying time originating from a lognormal distribution, which also provides a more smooth estimate over time.

The approach to estimate *R*(*t*) proposed by Khailaie et al. [2020] includes a complex SECIR-model with many assumptions about the other model parameters, which in part can only be guesstimated from literature sources. Their procedure of estimating *R*(*t*) is only partly data driven and mainly relies on cumulative reported cases in the federal states of Germany. Confidence intervals are generated by the variation of the other model parameters. This highlights the problems of the approach: While SECIR- models can be useful for forecasting, its value for real-time estimation of *R*(*t*) hinges on it being a realistic model with well calibrated parameter estimated. Instead, we prefer the more statistically driven transmission-tree based estimates, which rely less on model assumptions and more on a statistically sound analysis of the available data.

In our retrospective evaluation of the Bavarian COVID–19 data we found, that the estimation of *R_e_*(*t*) based on the predicted daily counts of newly diseased cases from nowcasting performs well if the nowcast model is adequately specified. Coverage frequencies of the 95% credibility intervals were as desired compared to a calculation of *R_e_*(*t*) based on all retrospectively available disease onset data. The utilization of the predictive distribution from the Bayesian nowcast for the estimation of *R_e_*(*t*) helps therefore successfully to avoid a bias close to the current date due to diseased-but-not-yet reported cases. For interpretation of the estimated 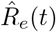 over time, similar limitations arise as in the interpretation of the estimated epidemic curve from the nowcast. If the fraction of undetected cases compared to all cases changes strongly over time, e.g., due to changes in testing strategy, this can bias the estimated reproduction number. Compared to the interpretation of the estimated epidemic curve, the time-dependent reproduction number might, however, have the advantage that it only requires stable conditions within a short time window, since it compares the estimated and reported number of newly diseased cases to the situation at time points close by, instead of looking at the absolute numbers over a longer period of time.

Summarizing, we believe that our results give a much more reliable picture of the course of the pandemic than the mostly used time series of reported cases. For the interpretation, it has to be emphasized, that we estimate the number of persons with disease onset on a certain day.

Our proposed nowcasting model can be applied to other data, when sufficient information about disease onset is available and the numbers are large enough for reliable modeling. On our webpage (corona.stat.uni-muenchen.de) we present daily results of the nowcasting for Bavaria and, in addition, for the city of Munich.

Since we introduce no correction for cases, which are never detected, our estimated epidemic curve should be related to other data sources, like hospital admission, ICU admission or death numbers. This aspect highlights the need for the collection and combination of many different data sources - each bringing challenges of its own.

## Data Availability

Code for reproducing our analysis and adapting it to other application scenarios is available at Github. There, we also provide an artificial dataset based on the observed reporting dates of cases but featuring only artificial information on the age and symptom onset dates of the cases.

https://github.com/FelixGuenther/nc_covid19_bavaria

## Acknowledgement

We thank Titus Laska for initial code on the *R_e_*(*t*) estimation. An early version of the imputation modeling, but without any methodological extensions of the nowcast, was proposed in the now disused work by Glöckner et al. [2020].

## Appendix

### A.1. Sensitivity analysis

Since the amount of data with missing information on disease/symptom onset is rather high, we perform two sensitivity analyses. The symptom onset date can either be missing because the reporting local health authorities were not able to provide any information or because a case did not experience any symptoms until case reporting. Based on available information, we can distinguish between cases for which COVID–19 symptoms are documented at time of reporting(17,723 (61%); 73% with available onset), cases explicitly without symptoms (2,221, 8%) and cases without any information on symptoms (9,302, 32%). In the first sensitivity analysis we focus only on the cases with reported symptoms. In a second analysis, we exclude all cases which have been explicitly reported to have no symptoms.

Figure 4 A indicates that the estimated structure of the epidemic curve is very similar to the main analysis when excluding asymptomatic cases and cases without known symptom status in the sensitivity analyses. This also applies to the estimated *R_e_*(*t*) as show in Figure 4 B. Since the sensitivity analyses consider fewer reported cases, the actual estimated number of new cases per day is lower as well. However, the interpretation regarding the dynamics of the COVID–19 pandemic in Bavaria is similar based on all three analyses.

